# Estimation of stratified seroprevalence directly from raw serological assay measurements with multi-level Bayesian mixture modelling

**DOI:** 10.64898/2026.09.17.26363301

**Authors:** Chris Wymant, Michelle Kendall, James A. Hay, Christophe Fraser

## Abstract

Estimating seroprevalence for an infection—the proportion of individuals with antibodies—and its variability between subpopulations is a key first step for research and the allocation of resources for treatment and prevention. The relevant raw data is often serological assay measurements that serve as a proxy for antibody level, such as optical density values from enzyme-linked immunosorbent assays (ELISA). Analysis pipelines typically proceed through sequential steps of fitting calibration data independently in each separate batch, transforming assay measurements to antibody levels via each fitted calibration relationship, classifying antibody levels into discrete serostatus by comparison to a threshold, and finally comparing seropositive proportions between subpopulations. Such approaches have numerous limitations including loss of information, overconfidence (discarding uncertainty), and bias from inappropriate choice of threshold. We developed a Bayesian statistical model that replaces the sequential steps of the typical pipeline by multiple levels within a single hierarchical model, treating each sample’s antibody level as a model parameter rather than directly observed data. This integrates the connections from the raw assay measurements all the way through to subpopulation variability in seroprevalence, allowing partial pooling of information between related variables and the propagation of uncertainty from each part of the model throughout the whole of the rest of the model. We calculate observation probabilities for all assay measurements from both samples and calibration data, allowing easy identification of outlying measurements. We replace a single hard classification threshold for disease status by continuous probabilities that are adapted to each subpopulation. We allow a flexible multivariate random-effects logistic regression to capture variability in seroprevalence between subpopulations. Using simulated data we found that the typical stepwise approach gave prevalence estimates far from the true values with narrow confidence intervals. Our method, *dvsb*, had markedly greater accuracy. We report the run time and convergence of *dvsb* when applied to a real dataset for Lassa fever IgG antibodies measured with ELISA, comprising 72,863 measurements for 21,391 unique samples (results reported elsewhere). *dvsb* is available at https://github.com/BDI-pathogens/dvsb.

## Introduction

Estimating the prevalence of a disease and its variability between subpopulations is a key first step in epidemiology, informing downstream study and the allocation of resources for treatment and prevention. For infectious diseases prevalence can be difficult to measure, and serological evidence of past infection can serve as a useful metric to understand epidemiological trends (Haselbeck et al. 2022; Hay et al. 2024). Seroprevalence is the proportion of individuals who are seropositive, i.e. with antibodies for the causative pathogen at a high enough level to suggest previous infection. Seroprevalence measures cumulative infections in the past, adjusted for the fact that some individuals who were previously seropositive may become seronegative by the time of observation. When coupled with an understanding of the protective effect provided by antibodies, seroprevalence also measures the population’s immunity, which can be important for forecasting disease dynamics and designing vaccine trials. Henceforth we will restrict our discussion to the estimation of seroprevalence for concreteness, but our arguments and method apply to the estimation of disease prevalence more generally.

Seroprevalence is measured using serological assays, which typically measure antibody levels indirectly, via proxy measurements such as optical densities in enzyme-linked immunosorbent assays (ELISA). A set of ‘calibrators’ with a range of known antibody levels are included on each ‘plate’ (or batch) of samples, allowing us to estimate the relationship between antibody levels and proxy measurements and how it varies by plate due to experimental conditions. This relationship then allows us to estimate the unknown antibody levels of samples from their proxy measurements. Here we focus on ELISA optical density (OD) measurements, but the same principles apply to other serological assays such as fluorescent intensities from multiplex bead assays and protein microarrays.

A typical analysis pipeline, starting from serological proxy measurements and ending with the variability in seroprevalence between subpopulations, proceeds stepwise as described in the first column of Table 1. This process has numerous limitations, described in the second column, some of which have been discussed previously (Kain et al. 2025). We refer back to each limitation at the start of Results.

**Table 1:** steps in a typical analysis pipeline from serological proxy measurements to variability in seroprevalence between subpopulations, and limitations of such an approach.

| Typical analysis pipeline step | Limitations |
| --- | --- |
| First, the relationship between proxy measurements, $y$, and antibody levels, $x$, is estimated for each plate within the dataset independently of all other plates, using only the calibration data for that plate in a homoscedastic regression model. | 1a. When analysing one plate, ignoring data from all other plates will cause overfitting to observational stochasticity on that plate. Plates are different from each other, but are not wholly independent of each other, under the assumption that their variability is describable by some distribution.<br>1b. The $y$ values observed for the samples on a plate can inform the estimation of the $x \leftrightarrow y$ relationship for the plate even though they lack an associated $x$ observation. |
|  | 1c. Homoscedasticity may be a poor approximation: the observational stochasticity (measurement noise) in $y$ may increase appreciably with $x$ (as we have found in ELISA data, not shown). |
| Second, each sample has a single antibody level estimated by transforming the sample's $y$ to $x$ using that plate's estimated relationship. If a single sample has multiple $y$ values from replicate measurements or retesting, a single $x$ value is obtained by taking a mean of $y$ values before transformation or a mean of $x$ values after transformation. | 2a. When estimating $x$ for one sample, ignoring data from all other samples will cause overfitting to observational stochasticity for that sample. Samples are different from each other, but are not wholly independent of each other, under the assumption that their variability is describable by some distribution.<br>2b. Uncertainty in the estimated $x \leftrightarrow y$ relationship should be propagated through into the $x$ values estimated for all samples.<br>2c. When multiple measurements of the same quantity are available, reducing them to a single value by taking a mean is unjustified when the process generating the variability between them has not been modelled, and may be inappropriate depending on what the process is. Furthermore there is no principled approach for testing the consistency between multiple measurements, which could reveal outliers/errors.<br>2d. If the common choice of a four-parameter logistic function is made for the $x \leftrightarrow y$ relationship, this has maximum and minimum possible $y$ values. Some samples may have $y$ values greater than the maximum that is estimated based on the calibration data, or less than the minimum. No $x$ value can be determined for such samples, and an ad hoc approach must be chosen such as manually setting their $x$ to zero or infinity. |
| Third, the continuum of $x$ values is binarised into seropositives and seronegatives by comparison to a single threshold, pre-specified for example by the manufacturer of the experimental kit or from an analysis of controls. | 3a. Uncertainty in the estimated $x$ value for each sample ought to be propagated through into its estimated serostatus.<br>3b. Manufacturer thresholds may be inappropriate: they are typically chosen based on some unknown optimisation of specificity or sensitivity, in a population perhaps differing from the one under analysis, leading to spectrum bias.<br>3c. The appropriate threshold to use depends on the subpopulation, being variable even within a single dataset (see Figure 1). |
| Fourth, counts of seropositives and seronegatives in each subpopulation define seroprevalence, and logistic regression with fixed effects is used to estimate which covariates define relevant variation in seroprevalence. | 4a. It is formally incorrect to classify each sample's serostatus first and estimate the subpopulation's seroprevalence second, because the first step depends on the second.<br>4b. For categorical predictor variables in a logistic regression, estimating the variation between categories assuming they are completely independent (i.e. using fixed effects) will overfit to observational stochasticity in each category. Categories are different from each other, but are not wholly independent of each other, under the assumption that their variability is describable by some distribution. |

Here we address all of the limitations of the typical pipeline listed in Table 1 by developing a statistical model for estimating seroprevalence and its variability between subpopulations directly from the proxy measurements obtained in serological assays. We use a single joint model for all data together, replacing the sequential steps of the typical pipeline by multiple levels inside a hierarchical model, allowing partial pooling of information between related variables and the propagation of uncertainty from each part of the model throughout the rest of the model. Calculating a probability for almost all aspects of the data allows easy identification of outlying measurements. We replace a single hard classification threshold for serostatus by continuous probabilities of being seropositive, adapted to each subpopulation following the logic of Bayes’ theorem. We describe the application of our method to simulated data, and comment on its application to real data reported elsewhere.

## Methods

### Bayes’ theorem for individual classification

The classic application of Bayes’ theorem to (serological) diagnostics tells us that the probability of being seropositive (‘pos’) rather than seronegative (‘neg’) given a measured antibody level x is

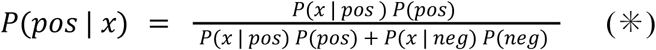

where P(pos) is seroprevalence, P(neg) is 1 - seroprevalence, P(x | pos) is the distribution for x among seropositive individuals, and P(x | neg) is the distribution for x among seronegative individuals. The denominator of the right-hand side is the distribution for x in the population comprising both seropositive and seronegative individuals in their true proportions; this is a ‘mixture’ distribution, made by mixing the distributions defined for seropositive and seronegative individuals separately. The whole fraction on the right-hand side can be described as the proportion of the mixture contributed by the seropositive component of it, evaluated at one specific value of x.

### Variation in Bayes’ theorem by subpopulation

If seroprevalence varies between subpopulations, then equation (✳) is adapted by conditioning every term in it on the subpopulation of interest, s say. P(pos | x, s) thus depends on s, and the threshold value of x that we choose for classification—for example, the value above which being seropositive is more likely, below which being seronegative is more likely—also varies with s. (One could choose other thresholds depending on the desired sensitivity-specificity trade-off, but dependence of the threshold on s remains.) In Figure 1, we illustrate this threshold variation for a toy population composed of two subpopulations which differ only in their seroprevalence: this alone causes the classification threshold to change. Subpopulations may differ in other regards that change the threshold, such as immunological or infection history differences shifting the mean x for seropositives.

**Figure 1:**
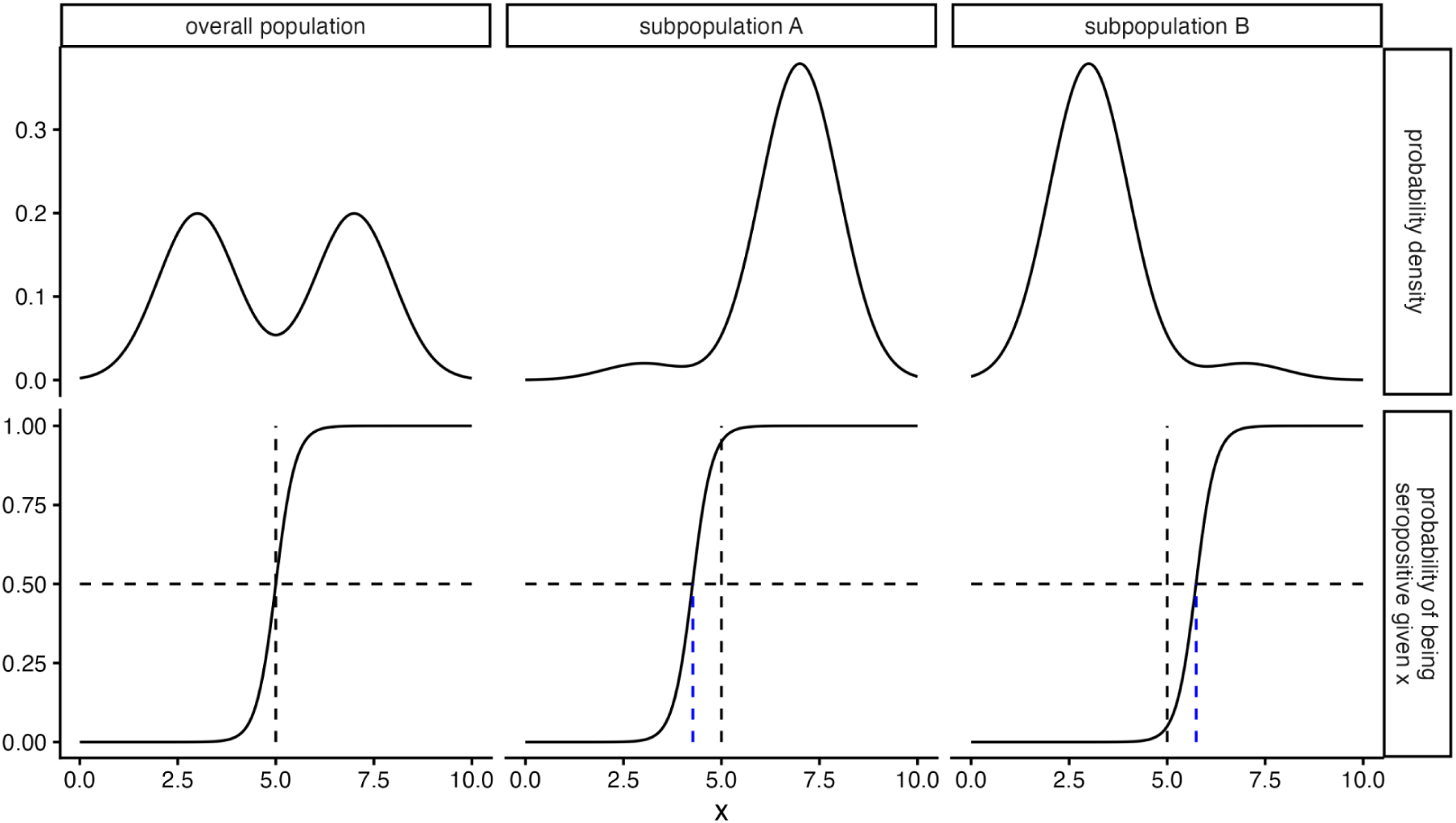
distributions for x (antibody level) and probabilities of being seropositive. We show results for a toy model in which an overall population (left column) is made up of 50% subpopulation A (middle column) and 50% subpopulation B (right column). Seroprevalence is 50% in the overall population, 95% in subpopulation A, and 5% in subpopulation B. Top row: distributions of x. Bottom row: the probability of being seropositive given x. The horizontal dashed line highlights a probability of 0.5: where the curve is right (left) of this, seropositive (seronegative) is the most likely disease status given x. Vertical dashed black lines show the value of x at which the probability of being seropositive is 0.5 in the overall population, i.e. the classification threshold. Vertical dashed blue lines show how that threshold shifts in the subpopulations. In the subpopulation plots, the difference between the blue and black dashed vertical lines show the difference between the correct and the naive (whole-population) classification thresholds, and the probability at which the black dashed horizontal line crosses the curve shows how much the seropositive probability has shifted away from 0.5 at the naive classification threshold.

### Seroprevalence comes first, individual classification second

The threshold for classifying an individual sample thus depends on the seroprevalence in that sample’s subpopulation. This might sound circular, because seroprevalence is usually defined by first classifying all samples and then finding the proportion classified as seropositive. However, we can estimate seroprevalence directly by estimating the distribution of antibody levels, decomposed into a contribution from seropositives and a contribution from seronegatives; the proportion of this mixture distribution that is contributed by seropositives equals the seroprevalence. Classification of individual samples comes second, following from this distribution, according to equation (✳). Estimating seroprevalence directly in this manner means we take into account uncertainty in each individual’s serostatus. To illustrate the usefulness of this logic, imagine we could estimate this distribution precisely, and we have a large number of samples all close to the classification threshold. We can be simultaneously highly uncertain of the status of each of these samples individually, yet highly certain that close to half of them are seropositive. By contrast, if attempting to classify individuals first, very little can be said about this set of samples.

### The hierarchy of variables

Figure 2 shows the relationship between the different levels of our multi-level statistical model. The rest of Methods describes each part of the model in turn using minimal equations; see Supplementary Information for a more precise description using equations. Top-level parameters (‘fixed effects’) have prior distributions, the parameters of which (‘hyperparameters’) are kept fixed during inference. Lower-level parameters (‘random effects’) have prior distributions that depend on other parameters that vary, i.e. parameters that form part of the parameter space we explore during inference. Top-level data is not modelled, rather it is conditioned upon; lower-level data is modelled. (For example in simple linear normal regression, P(y | x, m, c, σ) = N(y | mx + c, σ), both x and y are data but y is modelled whereas x is conditioned upon).

**Figure 2:**
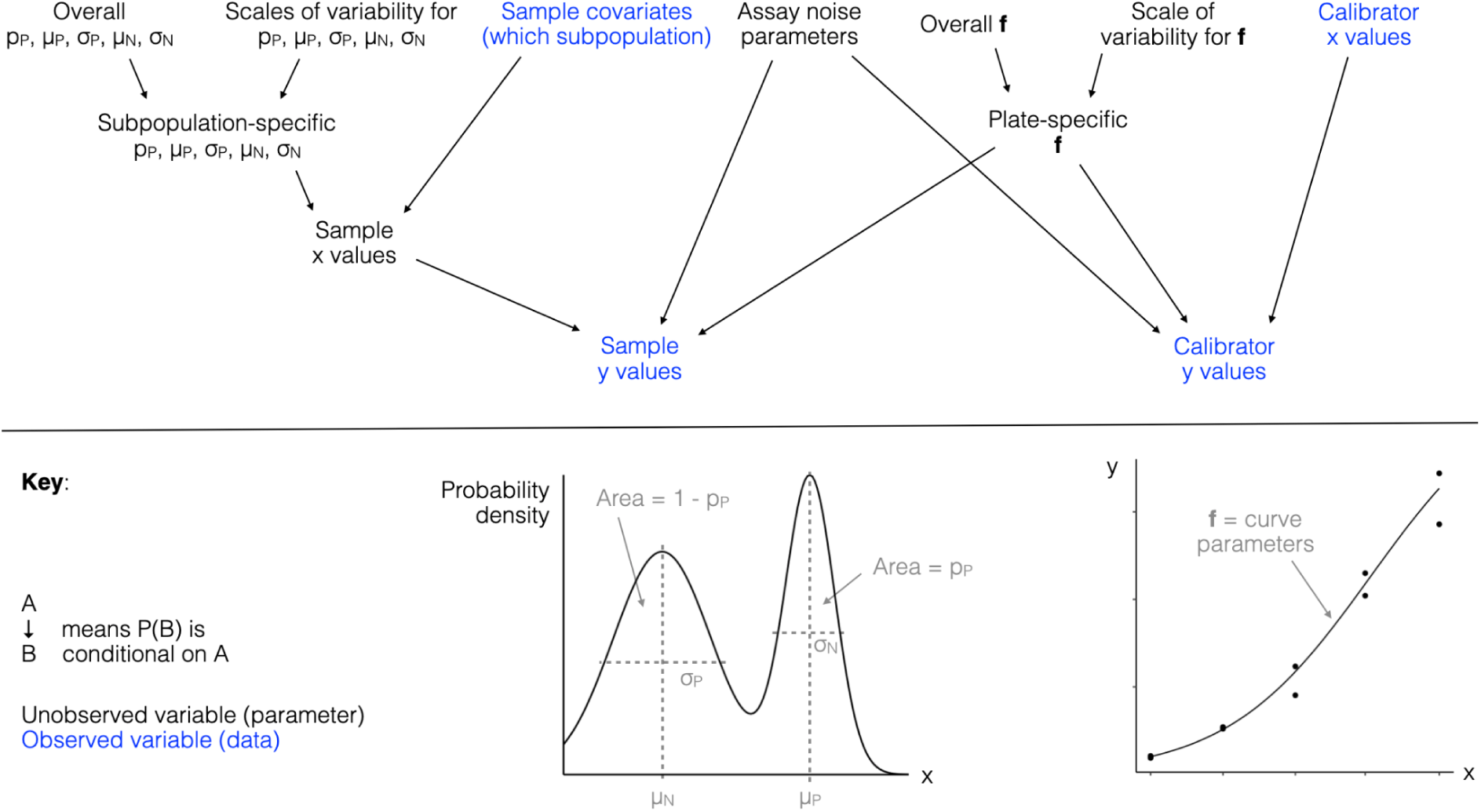
the relationship between the different levels of our multi-level statistical model. An arrow between variables indicates dependency in the probability distributions of those variables, i.e. the need to condition one variable’s distribution on the other variable. Abbreviations: x is antibody level, p_P_ is the proportion seropositive, μ_P_ and σ_P_ are the mean and standard deviation of x conditional on being seropositive, μ_N_ and σ_N_ are the mean and standard deviation of x conditional on being seronegative, **f** is the set of parameters describing the relationship between x and y (y is the proxy measurement reported by the assay, e.g. ELISA OD values).

### The basic mixture model

We model log antibody levels as having one normal distribution conditional on being seropositive, and another normal distribution conditional on being seronegative. The overall distribution—unconditional of serostatus—is thus a two-component normal mixture. More precisely, let x be the log antibody level in standardised units, p_P_ be the proportion seropositive, μ_P_ and σ_P_ be the mean and standard deviation of x conditional on being seropositive, μ_N_ and σ_N_ be the mean and standard deviation of x conditional on being seronegative. The distribution for x unconditional of serostatus is p_P_N(μ_P_, σ ^2^) + (1-p_P_)N(μ_N_, σ ^2^).

### Random effects on the mixture model

We model all five parameters of the two-component mixture distribution for x as varying between subpopulations. We use random effects to describe this variation, i.e. we partially pool estimates of these parameters between the subpopulations. Random effects on p_P_ are normally distributed on a logit scale (to ensure p_P_ always remains between 0 and 1). Random effects on σ_P_ and σ_N_ are normally distributed on a log scale (to ensure σ_P_ and σ_N_ are always positive). Random effects on μ_P_ and μ_N_ have a truncated normal distribution for identifiability (to break the seropositive↔seronegative re-labelling symmetry; see Supplementary Section 1.4). Subpopulations can be specified using multiple covariates simultaneously, and using different covariates for different parameters; for example, p_P_ could be set to vary by location and occupation, μ_P_ to vary by age group. When using multiple covariates for the same parameter, their combined effect is additive across covariates, and partial pooling only occurs for different categories of the same covariate. For example if a parameter is set to vary by location and occupation, we partially pool differences between locations and, independently of that, partially pool differences between occupations. In addition to the random effects, we allow one fixed effect on p_P_ for each of a series of binary predictor variables. Our specification of a regression model for p_P_, which is the probability of each sample’s latent binary serostatus parameter taking the value of seropositive, means our model is an example of ‘latent class regression’ (Kain et al. 2025). We do not model covariate data, we condition upon it (for example if age is used, we do not estimate the age distribution but take it as given).

### Relating x and y

Let y be the quantity directly measured by our serological assay, for example optical density in ELISA. We model the expected value of y as a function of x with some set of parameters **f**; specifically we use a four-parameter logistic (4PL) function. We model **f** as varying between ‘plates’ (subexperiments or batches within a larger dataset): there are plate-specific random effects on the x↔y relationship. Specifically, **f** follows a four-dimensional normal distribution. A regression model can also be specified for systematic differences in **f** between groups of plates. We model the observational stochasticity of y around its expected value as normally distributed with a variable standard deviation, i.e. allowing heteroscedasticity. Specifically, we use another 4PL function for this standard deviation, sharing two of the parameters from the 4PL for the expected value of y (those for the location and steepness of the jump) but introducing two new parameters for the scale of observational stochasticity at asymptotically small or large x specifically for calibration data, and two equivalent parameters for samples (which we found in our data to have more stochastic y than calibrators do). Our model for the relationship between x and y thus captures that larger x results in larger y but also larger stochasticity in y, with both aspects having a sigmoidal shape. The calibration data is important in informing this relationship, but sample data also contributes. We model calibrator x values as exactly equal to some a priori known value.

### Accidental blanks

Our method includes an extension of the main statistical model that allows for the possibility of, and estimates the frequency of, some wells in plates accidentally containing no sample at all. In this model the distribution for y for a given sample is not a pure normal whose mean and standard deviation are 4PL functions of that sample’s antibody level, as described above. Instead it is a mixture of that normal and a second normal whose mean and standard deviation are appropriate to an antibody level of zero. We do not use this alternative model for the results presented here.

### Posterior retrodictive checks in our model

Reliable statistical modelling requires checking model fit, i.e. how well the data is modelled, for example by posterior retrodictive checks. We facilitate and recommend three posterior retrodictive checks in our method. The first is for the y values for each calibrator across all plates given their x values, to check how well we model the x↔y relationship across all plates. The second is for the set of y values from each sample, to check for any inconsistencies between multiple measurements from the same sample, and for any samples with extreme y values that are in tension with the assumed underlying distribution for x given our estimated x↔y relationship. The third is the subpopulation-level distribution of y values over all distinct subpopulations that are modelled as having different x distributions. Assuming the x↔y relationship has been modelled well, this checks how well we model the distribution of x and its variability between subpopulations.

### Parameter recovery testing

We tested the correctness of our implementation of our statistical model by recovering known parameters values used to simulate data, using the same generative process for simulation as in the inference model. For this we simulated datasets with a variable number of plates, each plate containing 40 samples plus 6 calibrators (including a blank), with two observations of each sample and calibrator (measurements in duplicate). We simulated three subpopulations each differing in their means and standard deviations for x for both seronegatives and seropositives. Assay parameters were set to roughly match those we observed in real IgG ELISA data for Lassa fever (ENABLE Consortium, in prep.).

### Comparison to the typical pipeline

We compared the performance of our method to that of the more typical analysis pipeline described in the first column of Table 1. For this we simulated data from a population composed of two equally sized subpopulations with one having twice the seroprevalence of the other, allowing us to test estimation of two seroprevalences and of their difference. We varied the overall seroprevalence through values of 1%, 10% and 50%; this controlled each individual’s probability of being seropositive, such that the actual fraction of positive individuals in each stochastic replicate of dataset simulation varied slightly around these values. We simulated a sampled population of 1000 individuals for 10% and 50% overall seroprevalence, and 5000 individuals for 1% overall seroprevalence, repeating the simulation five times for each scenario analysed using a different seed for random number generation. For the typical analysis pipeline, first we estimated the maximum-likelihood 4PL x→y relationship independently for each plate using only the calibration data, using homoscedastic regression from the dr4pl R package (Landis et al. 2021); second, we took the mean of each sample’s duplicate y values and transformed this into one x value by inverting the x→y relationship; third, we classified each sample as seropositive or seronegative based on a single threshold defined as some multiple number of standard deviations greater than the seronegative mean (assuming this is correctly known); fourth, we estimated seroprevalence in each of the two groups with binomial confidence intervals, and estimated the difference in seroprevalence on a logit scale using logistic regression.

### Implementation

We implemented our statistical model as code in the Stan language (Carpenter et al. 2017) (Stan Development Team 2026b), interfaced with one of rstan (Stan Development Team 2026a), cmdstanr (Jonah Gabr, Rok Češnova, Andrew Johnson, Steve Bronder 2025), or cmdstan, as selected by the user. R code controls input and analysis of output. We named our method *dvsb*: Disease Variability in Subpopulations from Biomarkers. The code is publicly available at https://github.com/BDI-pathogens/dvsb.

### Application to real data

For our application to real data, we partitioned the full dataset into seven geographical sites, and ran *dvsb* independently on each partition. We preferred this to a single run on all data simultaneously, firstly for computational pragmatism given the dataset size, and secondly because this allowed the flexibility of every model parameter varying by site (each site had enough data that pooling added negligible benefit, and some but not all parameters of the model are allowed to vary by subpopulation within a single analysis). We ran our method using four chains multithreaded on four cores (i.e. one core per chain), on the University of Oxford Biomedical Research Computing cluster. The runtime we report is the time taken by the slowest of the four chains. The R hat we report is the largest R hat (Vehtari et al. 2021) of any parameter.

## Results

### How traditional limitations are solved

We begin by clarifying how our method *dvsb* addresses the limitations of the traditional analytical pipeline from Table 1. First, for limitations 3b, 3c and 4a, we use a mixture model (varying by subpopulation) to distinguish seropositives from seronegatives, and in doing so we adjust the threshold to the (sub)population analysed. Mixture models are the most appropriate way to estimate seroprevalence when many seropositive and many seronegative samples are available, without pre-existing serostatus labels (White et al. 2026). Second, for limitations 2b and 3a, we use a single multi-level model instead of a sequence of steps, and in doing so we propagate uncertainty in each part of the analysis through into the rest of the analysis. This is important not only for correctly caveating results with sufficient uncertainty, but also for accuracy: treating every sample even slightly below (above) some manufacturer threshold as definitely negative (positive, respectively) can make seroprevalence estimates inaccurate. Third, for limitations 1a and 2a, by jointly modelling all samples and plates together—estimating their individual properties together with the distributions that characterise their variability—we reduce overfitting to stochastic noise in individual samples and plates. Fourth, for limitation 1b, we use samples, not only calibrators, to inform estimation of the relationship between antibody levels (x) and the serological assay proxy values (y) on each plate. This is possible even without knowing the x of each sample, because we do know that multiple replicates of the same sample, on the same plate or different plates, have the same x. Fifth, for limitations 1c, 2c and 2d, we model the process generating variability between y values from multiple replicates of the same sample starting from a single x, and in doing so we avoid the need to take a mean of multiple y values to get back to a single x. Taking a mean here is not only unprincipled but inaccurate, given the nonlinear heteroscedastic relationship we use to model both the expectation and the noise in y to the underlying x. For example with one large and one small y, the larger one carries more weight: a stochastically small y from a large x is more likely than a stochastically large y from a small x. An additional benefit to modelling this process is being able to calculate each replicate’s probability under the model, facilitating identification of outlying measurements from experimental problems. Furthermore, since we model the generative process forwards from a sample’s x to its y, we avoid the aforementioned problem of impossible y values (greater than the maximum or less than the minimum) that arises when attempting to calculate x from y by simple inversion of the relationship. Sixth, for limitation 4b, we use random effects rather than fixed effects to distinguish the effect of different population categories, such as of age groups or places on seroprevalence, and in doing so we reduce overfitting to stochastic noise in each category. This pulls estimates from small groups with outlying results towards the better-characterised groups, by an amount determined by the scale of variability between the groups.

### Parameter recovery testing

We tested the correctness of our implementation of our statistical model by recovering known parameters values used to simulate data. Figure 3 shows the prior distribution, posterior distribution and true value for a subset of parameters. As the size of the dataset grows, the posterior becomes increasingly concentrated at the true value. The model fit to the distribution for y also becomes concentrated at the empirical distribution (Supplementary Figure 1).

**Figure 3:**
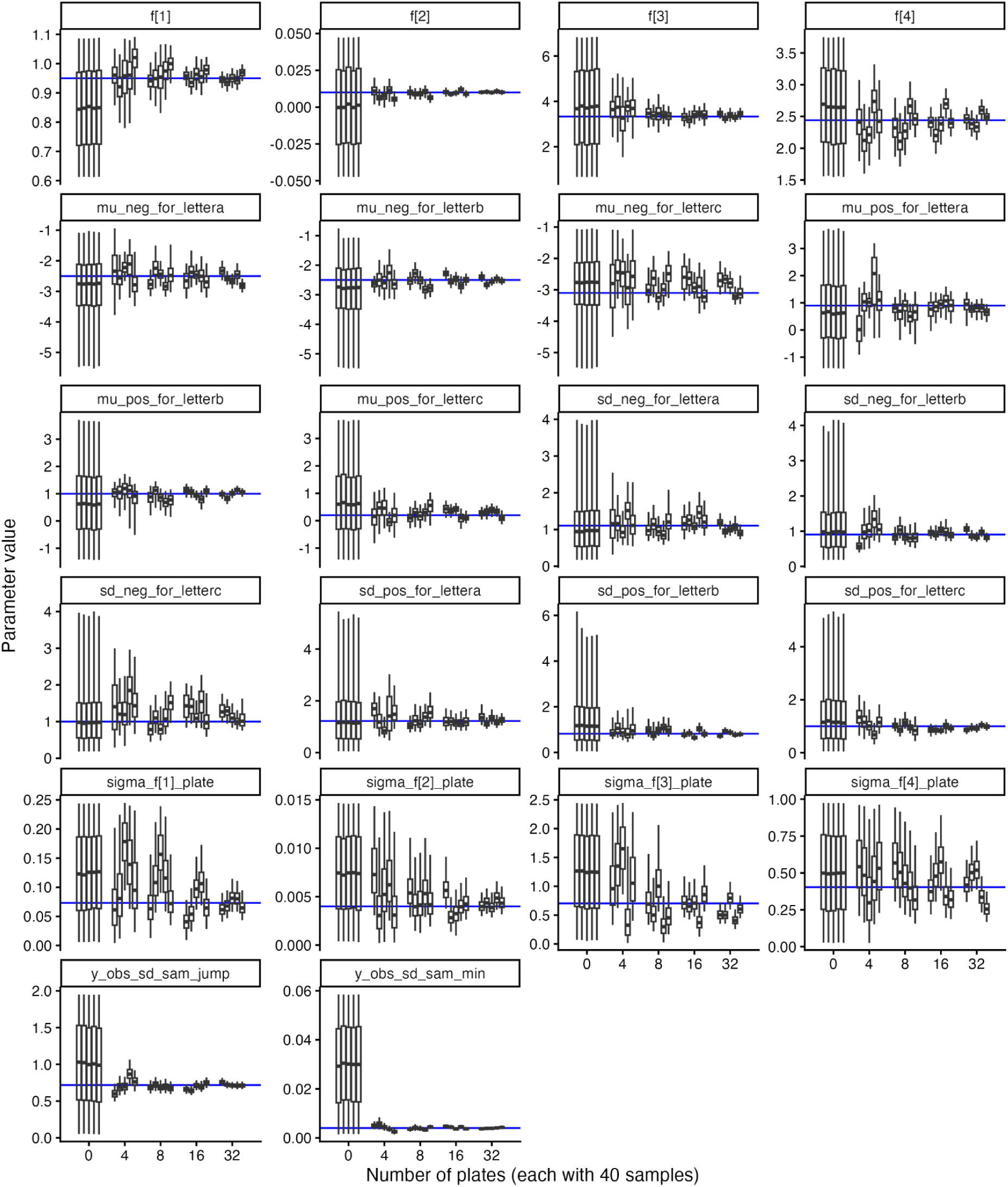
results from an application of our method to simulated datasets, showing posterior estimates of a subset of parameters. The x axis shows variation in the size of the dataset (for zero data the posterior equals the prior). The black horizontal line, box and whiskers show the posterior median, interquartile range and central 95% respectively; five of these are shown for each dataset size, for different stochastic replications of dataset stimulation. The blue horizontal line shows the true parameter value used for simulation. f[1-4] are the four parameters of the logistic function relating x to y. sigma_f[1-4]_plate are the scales of variability in f[1-4] between plates. mu_neg, mu_pos, sd_neg, sd_pos are the means and standard deviations for seronegatives and seropositives; the suffix ‘_for_lettera’ denotes the value of one of those four parameters for subpopulation a (and similar for b and c). y_obs_sd_sam_min is the scale of samples’ observational noise in y at asymptotically small x; y_obs_sd_sam_jump is the increase in that scale at asymptotically large x.

### Comparison to the typical pipeline

Using simulated data we compared the performance of *dvsb* to that of the more typical stepwise analysis pipeline described in the first column of Table 1. We varied how much greater the mean x is for seropositives than for seronegatives, which affects the overlap in the resulting distribution for y and the performance of all methods. For the typical pipeline we varied the threshold used to classify serostatus, which affects the bias in the estimation. Three or four standard deviations above the seronegative mean have been widely used as thresholds (Kain et al. 2025); we show results for two, three and four standard deviations. Figures 4 and 5 show results for a population with an overall seroprevalence of 50% and 1% respectively, made up of two equally sized subpopulations whose seroprevalences differ by a factor of two. For estimating the difference in seroprevalence between the two subpopulations (on a logit scale, i.e. the difference in their log odds of seropositivity), the stepwise method usually gave reasonable results. However, for estimating the seroprevalence in each group separately, the stepwise method gave estimates far from the true values with narrow confidence intervals: overconfident inaccurate estimation. The extent and the direction of the problem depended on the overall seroprevalence, the degree of separation in the distributions for x between seropositives and seronegatives, and the classification threshold chosen to distinguish seropositives from seronegatives. At 50% seroprevalence, the stepwise method overestimated seroprevalence unless the seropositive and seronegative distributions for x were strongly separated. At 1% seroprevalence, the stepwise method with a two-sigma classification threshold always overestimated seroprevalence; with a greater (stricter) classification, it underestimated seroprevalence unless the seropositive and seronegative distributions for x were well separated. Estimations from *dvsb* were markedly more accurate across the scenarios analysed. We show results for an overall seroprevalence of 10% in Supplementary Figure 2.

**Figure 4:**
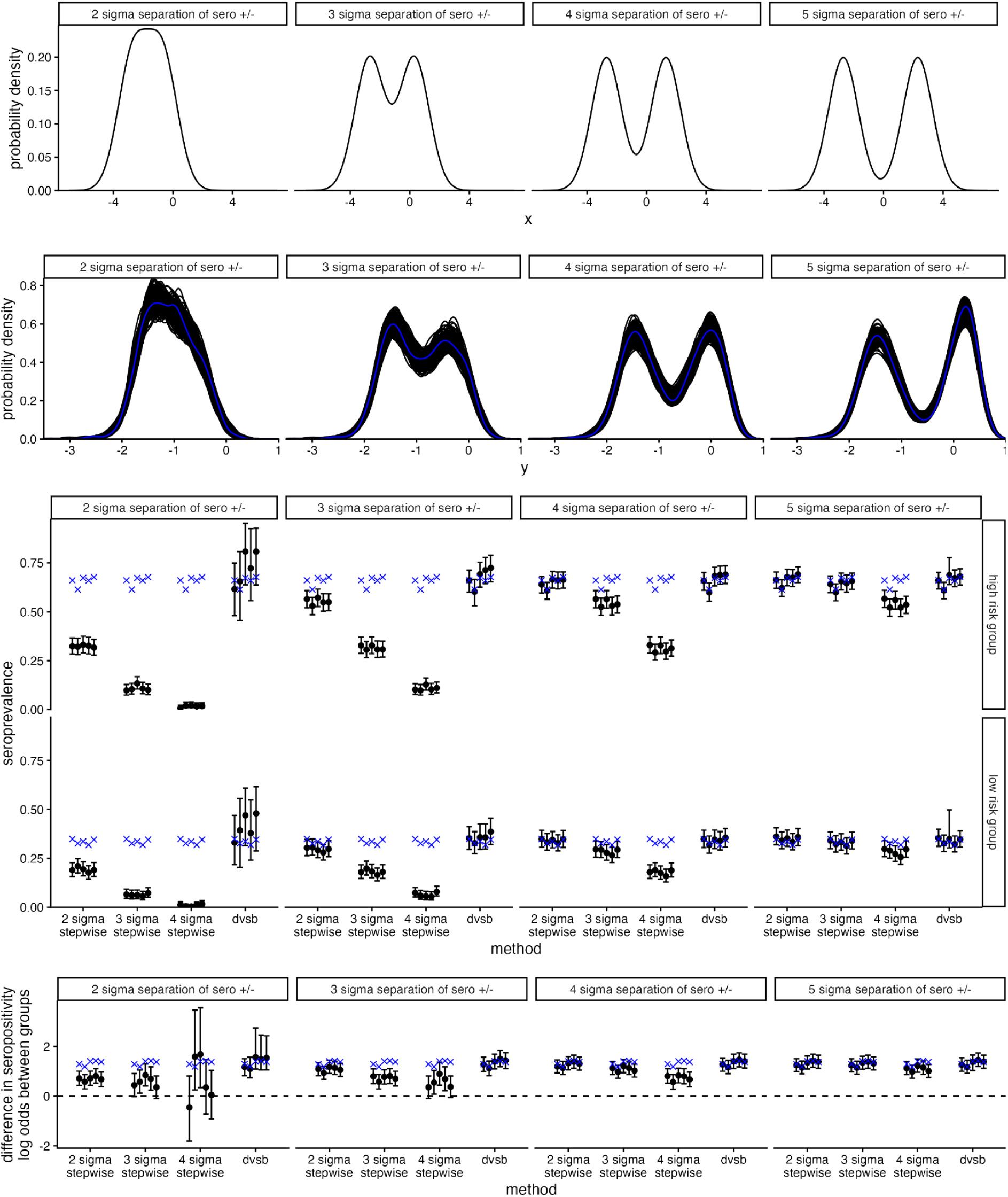
results for a simulated population composed of two subpopulations differing in their seroprevalence, with an overall seroprevalence of 50%. Each column shows a different level of separation between the x (antibody level) distribution for seropositives and that for seronegatives, e.g. “2 sigma separation of sero +/-” denotes that the mean x for seropositives is 2 standard deviations above the mean x for seronegatives (with both having the same standard deviation). First row: the overall distribution for x (including both subpopulations). Second row: the distribution for y (the serological assay proxy measurement for x, such as ELISA OD value), as a kernel density estimate, showing the empirical distribution in blue and the posterior fit to this distribution in black (one line per sample from the posterior). We exclude the small fraction of negative y values to allow a logarithmic scale for clarity. Third row: seroprevalence in the high risk subpopulation (upper third row) and low risk subpopulation (lower third row). Estimates are shown for our method *dvsb*, and for the typical stepwise method of analysing such data using different thresholds for classifying serostatus, e.g. “2 sigma” denotes classifying as seropositive all samples with an estimated x that is at least 2 standard deviations above the mean x for seronegatives. The black points and vertical bars show the posterior median and central 95% interval for *dvsb*, and the maximum-likelihood estimate and 95% confidence interval for the stepwise approach. Different points and bars for the same method show different stochastic replications of dataset stimulation. Blue crosses show the true seroprevalence for the population as stochastically sampled (which varies slightly between dataset replicates). Fourth row: the estimated difference in seroprevalence between the two subpopulations, on a logit scale. Blue crosses show the true difference. The horizontal dashed black line shows a value of zero for reference, i.e. no difference.

**Figure 5:**
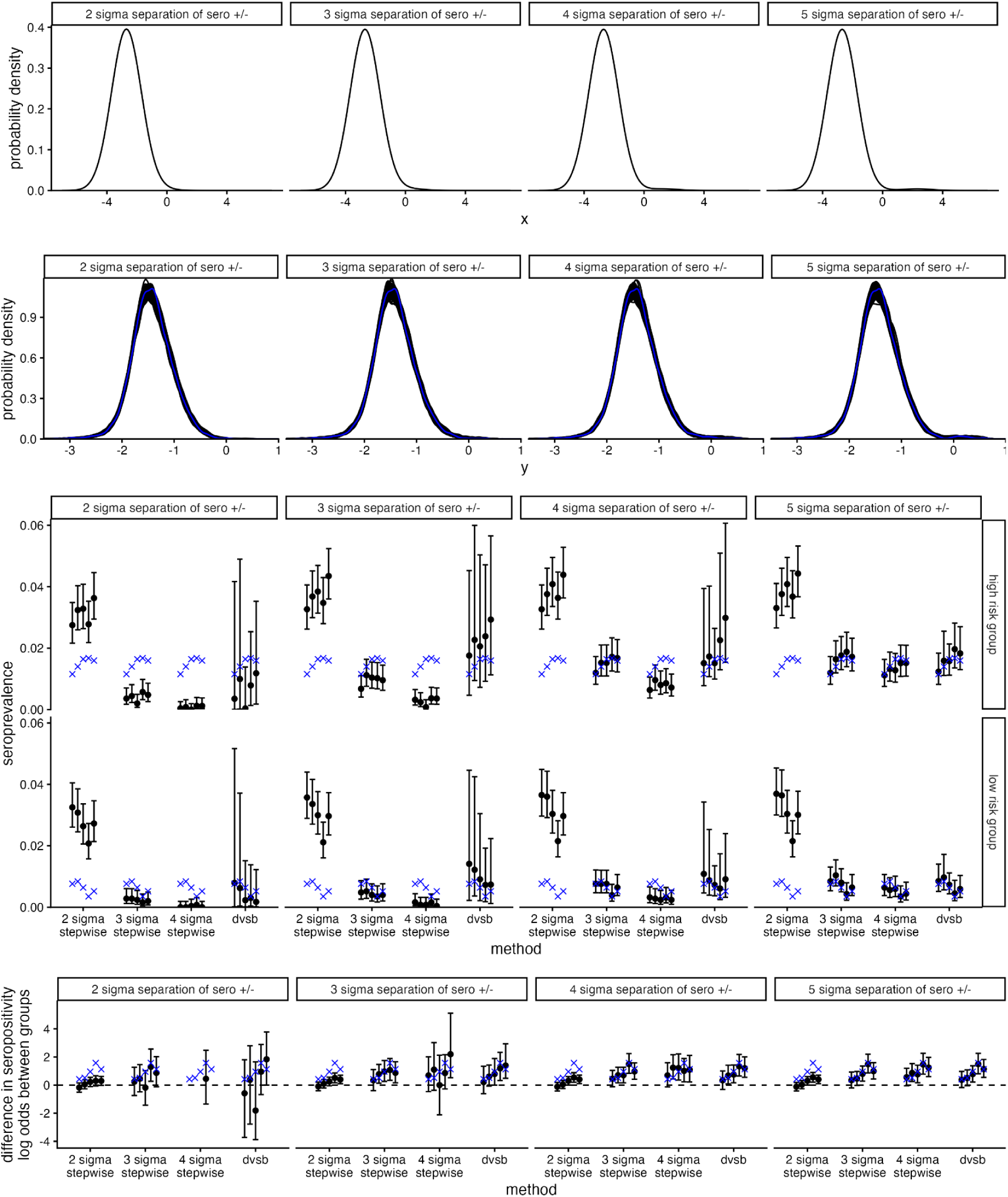
as Figure 4 but with an overall seroprevalence of 1%. In the fourth row, “2 sigma separation of sero +/-” plot, “4 sigma stepwise” category, four of the five estimated points are not shown due to having values dramatically outside of the range of the other points (highly inaccurate estimation).

### Application to real data

Elsewhere we describe results from an application of *dvsb* to real IgG ELISA data for Lassa fever in West Africa (ENABLE Consortium, in prep.). In Table 2 we provide summary metrics about the application itself (not the results emerging from it), to give a sense of how *dvsb* scales with the size of real data. For five of the sites, the extent of model convergence we obtained required roughly 2-5 hours per thousand unique samples. The other two sites were noticeably slower to converge; one reason could be greater difficulty in reconciling results from several different plates for a single sample (which we model as all sharing a single underlying x value, though in reality there may be differences due to variable sample storage and preparation, and in rare cases mislabelling).

**Table 2:** summary metrics about the application of our method *dvsb* to real IgG ELISA data for Lassa fever in West Africa.

| Site | Number of unique samples | Number of sample replicates | Number of calibrator replicates | Number of plates | Runtime (hours) | R hat |
| --- | --- | --- | --- | --- | --- | --- |
| 1 | 975 | 3197 | 2303 | 194 | 4 | 1.003 |
| 2 | 997 | 3590 | 2124 | 179 | 5 | 1.007 |
| 3 | 1002 | 3310 | 1366 | 114 | 54 | 1.025 |
| 4 | 4048 | 8263 | 2143 | 179 | 7 | 1.008 |
| 5 | 4589 | 12036 | 3888 | 326 | 24 | 1.001 |
| 6 | 4876 | 11387 | 3047 | 257 | 17 | 1.054 |
| 7 | 4904 | 12251 | 3958 | 334 | 15 | 1.003 |

## Discussion

In summary we developed a method, *dvsb*, for estimating disease seroprevalence and its variability between subpopulations directly from raw uncalibrated measurements from serological assays. We have focussed on seroprevalence for concreteness, but our arguments and method apply to biomarker-based estimates of disease prevalence more generally. Standard analysis pipelines proceed in a stepwise manner from serological assay proxy measurements (such as ELISA OD values), to point estimates of antibody concentration, to point estimates of serostatus, to logistic regression point estimates and confidence intervals for differing seroprevalences across subpopulations. By using a multi-level Bayesian mixture model to jointly model all data, we addressed many limitations of such pipelines, notably of their overconfident inaccurate estimation of seroprevalence. We demonstrated markedly more accurate estimation using *dvsb*.

Our work builds on a growing literature demonstrating the limitations and pitfalls of binary disease state classification using biomarker measurement thresholds (White et al. 2026; Chan et al. 2021). Numerous studies have demonstrated the added value of using mixture models or latent class analysis to estimate seroprevalence and serostatus without using a pre-specified cutoff (Kafatos et al. 2016; Bouman et al. 2021; Bottomley et al. 2021). Some used mixture models to update previously defined cutoffs, aiming to define subgroup-specific thresholds optimised for predictive accuracy (Tessier et al. 2023; Yang and Laven 2022; Parker et al. 1990; Yman et al. 2026). Others argue for avoiding binary classification entirely, propagating uncertainty in an individual’s state through downstream analysis, and avoiding misclassification of ambiguous measurements near the cutoff (often termed indeterminant or equivocal samples) (Kain et al. 2025). We argue for the latter approach, where serostatus should be treated as a probability rather than binary classification where possible; however, we note the practical advantages of binary classification, for example when clinical decisions need to be made for individual patients. Such threshold derivation is still possible using our approach. A benefit of the hierarchical modelling approach taken here is the inclusion of covariates into the estimation of seroprevalence and mixture component parameters as shown previously (Kain et al. 2025; Pfeiffer et al. 2000; Glemain et al. 2024).

A key novelty of our method is to jointly model the observation process of the serological assay itself alongside the mixture model and seroprevalence, treating each sample’s antibody level as a model parameter to be estimated with uncertainty, instead of as directly observed data. Previous work has used hierarchical models to adjust for batch effects and measurement error from raw serological assay data (Wang et al. 2026; Gelman et al. 2004; Swart et al. 2021) but ours is the first, to our knowledge, to propagate uncertainty in this process through to serostatus and seroprevalence estimates. While our method was developed for ELISA OD measurements, it could be extended to other assay types where the observation is a proxy for the underlying quantity of interest (antibody level), such as multiplex bead assay fluorescence intensity (FI), or plaque or foci reduction neutralisation test counts.

Our method has limitations. First, we model serostatus as having only two possibilities: seronegative and seropositive. More nuanced modelling could further distinguish categories by number of previous infections, or by cross-reactivity with other pathogens (Yman et al. 2026) (O’Driscoll et al. 2025). Second, for pragmatism, we have not modelled interaction effects between the different predictor variables for the parameters of the antibody level mixture model. Considering the example of age groups and spatial groups, this means that we model the difference in seroprevalence (on a logit scale) between two age groups as the same across all spatial groups. Third, we modelled antibody levels as having a (log) normal distribution; we modelled the relationship between antibody level and expected proxy measurement as a four-parameter logistic function; we modelled observational stochasticity, i.e. the difference between expected and observed proxy measurements, as normally distributed (with variable variance). These functional forms were sufficiently flexible to describe the real dataset that motivated our development of *dvsb*: ELISA data for Lassa fever. However, for other applications with different assays or diseases, different functional forms may be needed. The inclusion of samples with known serostatus (which were not available in our motivating data) could also benefit other applications. These features could be added to our code in future updates. Fourth, we have not modelled continuous predictor variables. Estimating seroprevalence as a continuous function of age lies at the heart of serocatalytic models, and the waning of antibody level with time since previous infection is best modelled as a continuous process. We hope to return to this in future work. Fifth, we use a two-component normal distribution to model the generation of log antibody levels for seronegatives and for seropositives; one of the two components will dominate at both asymptotically high and asymptotically low values. We elaborate on this issue in Supplementary Information section 1.5.

The takeaway from our study is our recommendation to estimate disease prevalence and its variability between subpopulations using *dvsb*, or similar multi-level statistical models that connect the raw observations through to the desired estimands within a single model. Compared to traditional stepwise analysis pipelines this improves estimation. Better understanding of subpopulation-stratified disease risk should improve downstream study of disease, trial planning, and the allocation of resources for treatment and prevention.

## AI

We used no artificial intelligence for writing code or this manuscript (excluding automatic spelling and grammar checks).

## Data Availability

Only simulated data are presented

## Acknowledgements

We thank Anton Camacho, Robert Hinch and Lucie Abeler-Dörner for helpful conversations. This project was funded by CEPI for the PADOVAX ChAdOx1 Lassa Project, PRJ-6057. JAH is supported by a Wellcome Trust Early Career Award (225001/Z/22/Z). JAH has received consulting fees from Gerson Lehrman Group (GLG).

