## Supplementary Information for "Estimation of stratified seroprevalence directly from raw serological assay measurements with multi-level Bayesian mixture modelling"

### 1 Mathematical definition of the statistical model

#### 1.1 Mathematical notation

We use standard mathematical notation, summarised below.

- $:=$ means 'is equal to by definition', serving to define the quantity appearing on its left.
- Bold font distinguishes vector or tensor quantities (when at least one of their indices is suppressed) from scalar ones. e.g. for matrix $\mathbf{M}$ and vector $\mathbf{v}$, $\mathbf{Mv}$ is a vector whose $i$th element is $\sum_j M_{ij} v_j$.
- As the index of a tensor quantity, a dot denotes taking the collection of all values of the tensor from all values of that index. e.g. if $\mathbf{X}$ is a matrix with elements $X_{ij}$, then $\mathbf{X}_{\cdot j}$ denotes the $j$th column of $\mathbf{X}$ (which is a vector), and $\mathbf{X}_{i\cdot}$ denotes the $i$th row of $\mathbf{X}$ (also a vector). We do not distinguish between row vectors and column vectors.
- $P(\ldots)$ means the probability of something. Also, with the common abuse of notation, the probability density of something.
- $\mid$ means 'given that' or 'conditional upon'.
- $x \sim f(\theta)$ means $x$ follows distribution $f$ parameterised by $\theta$. If $x$ is discrete this is a probability mass distribution; if $x$ is continuous this is a probability density distribution.
- $f(x \mid \theta)$ means the aforementioned distribution evaluated at $x$, i.e. $P(x \mid \theta) = f(x \mid \theta)$.
- A 'boolean' variable is one that must equal either 0 or 1.
- $s \sim \text{Ber}(p)$ means the boolean variable $s$ for the number of successes from a single trial is Bernoulli distributed with success probability $p$.
- $x \sim \text{Unif}(a, b)$ means scalar $x$ is uniformly distributed between lower limit $a$ and upper limit $b$.
- $x \sim N(\mu, \sigma^2)$ means scalar $x$ is normally distributed with mean $\mu$ and variance $\sigma^2$.
- $x \sim N(\mu, \sigma^2)[a, b]$ means scalar $x$ has a truncated normal distribution with mean $\mu$, variance $\sigma^2$, and support $[a, b]$ (i.e. $x$ is restricted to the range $[a, b]$).

- $\mathbf{x} \sim N(\boldsymbol{\mu}, \boldsymbol{\Sigma})$ means vector $\mathbf{x}$ is multivariate-normally distributed with mean $\boldsymbol{\mu}$ and covariance matrix $\boldsymbol{\Sigma}$.
- $\text{logit}(p) := \ln(p/(1-p))$, converting probabilities to log odds.
- $\text{logistic}(o) := \exp(o/(1+o)) = \text{logit}^{-1}(o)$, converting log odds to probabilities (the inverse of the logit function)
- $\text{4PL}(x \mid \mathbf{f}) := f_2 + (f_3 - f_2)/(1 + \exp(-f_1(x - f_4)))$ is the 'four-parameter logistic' function, mapping from the domain $[-\infty, \infty]$ to the range $[f_2, f_3]$, using four parameters collected into the vector $\mathbf{f} = (f_1, f_2, f_3, f_4)$
- Curly braces $\{\}$ denote a set (a collection of objects with no duplicates, with no concept of ordering). Caligraphic typesetting (e.g. $\mathcal{S}$ instead of $S$) indicates that a variable is a set. For a set the 'absolute' operator, $|\ldots|$, means the number of elements in the set.
- $\in$ means 'in', or 'is an element of'. Used for sets, e.g. if set $\mathcal{S} = \{1, 2, 3\}$, then $1 \in \mathcal{S}$ is true.
- $\cup$ means 'the union of'. $\mathcal{A} \cup \mathcal{B}$ is the set consisting of all elements of $\mathcal{A}$ and all elements of $\mathcal{B}$.

### 1.2 Definitions of variables

- $\mathcal{I} = \{1, 2, \ldots I\}$ is the set of all $I$ individuals in the study.
- $i \in \mathcal{I}$ indexes the individual, or equivalently the sample, since we consider exactly one sample per individual (though the same sample can be tested multiple times). Following common abuse of notation, we think of $i$ either as being an integer denoting which individual we are talking about, or as being an actual individual.
- $\mathcal{C} = \{1, 2, \ldots C\}$ is the set of all distinct $C$ calibrators used in the study (considering the same dilution on different plates to be distinct calibrators).
- $c \in \mathcal{C}$ indexes the calibrator.
- $\mathcal{L} = \{1, 2, \ldots L\}$ is the set of all $L$ plates in the study.
- $l \in \mathcal{L}$ indexes the plate.
- $\mathcal{R} = \{1, 2, \ldots R\}$ is the set of all $R$ replicates of a given observation (sample or calibrator) on a given plate. For example, if every sample and calibrator is measured in duplicate, $R$ is always 2.
- $r \in \mathcal{R}$ indexes the replicate.
- $s_i = \begin{cases} 1, & \text{if } i \text{ was seropositive at time of sampling,} \\ 0, & \text{if } i \text{ was seronegative at time of sampling} \end{cases}$
- $x$ is $\log_e$ of the antibody level (in some measurement unit that is used consistently for every observation).
- $y$ is the optical density (OD) value observed with the ELISA assay: the difference in absorption at the two wavelengths used. Specifically, $y^{\text{sam}}_{ilr}$ is the OD value of sample $i$, on plate $l$, replicate $r$; $y^{\text{cal}}_{clr}$ is the OD value of calibrator $c$, on plate $l$, replicate $r$. (These are only defined if individual $i$ or calibrator $c$ was present on plate $l$.)
- $\mathbf{f}_{l\cdot}$, the $l$th row of matrix $\mathbf{f}$, is the vector of parameters of the relationship between y and x on plate $l$.

- $\mathcal{V} = \{\mathcal{V}_1, \mathcal{V}_2, \ldots \mathcal{V}_V\}$ is the set of all $V$ sets (i.e. categorical variables) that are used as covariates to predict at least one of the five parameters of the two-component normal mixture model for log antibody levels ($p^{\text{pos}}, \mu^{\text{pos}}, \mu^{\text{neg}}, \sigma^{\text{pos}}, \sigma^{\text{neg}}$; see section 1.3). $\mathcal{V}' \in \mathcal{V}$ indexes the set within the set of sets, and $v \in \mathcal{V}'$ indexes the element therein. Each $\mathcal{V}'$ in $\mathcal{V}$ is a set of categories that one particular categorical predictor variable can be equal to. For example we might have $\mathcal{V}_1$ as a set of countries (with $v \in \mathcal{V}_1$ being one of the $|\mathcal{V}_1|$ countries) and $\mathcal{V}_2$ as a set of age groups (with $v \in \mathcal{V}_2$ being one of the $|\mathcal{V}_2|$ age groups). The same as for the abuse of notation mentioned above for the index $i$, we consider $\mathcal{V}'$ as either a set or as an integer index for which set we are talking about. Thus we use $\mathcal{V}'$ as an index appearing in subscripts and we consider which elements are in the set $\mathcal{V}'$.
  - $\mathcal{V}^\theta$ is the subset of $\mathcal{V}$ consisting of only those sets used as covariates to predict $\theta$, where $\theta \in \{p^{\text{pos}}, \mu^{\text{pos}}, \mu^{\text{neg}}, \sigma^{\text{pos}}, \sigma^{\text{neg}}\}$
  - Therefore we have $\mathcal{V} = \mathcal{V}^{p^{\text{pos}}} \cup \mathcal{V}^{\mu^{\text{pos}}} \cup \mathcal{V}^{\mu^{\text{neg}}} \cup \mathcal{V}^{\sigma^{\text{pos}}} \cup \mathcal{V}^{\sigma^{\text{pos}}}$
  - These definitions may become easier to understand once these quantities are seen in action, in equations 10–14.
- $X_{i\mathcal{V}'v} = \begin{cases} 1, & \text{if } i\text{'s category for predictor variable } \mathcal{V}' \text{ is } v, \\ 0, & \text{otherwise} \end{cases}$
  Instead of the more usual design matrix, this defines a 3D design tensor $\mathbf{X}$ that is ragged: there are different numbers of $v$ indices for different values of $\mathcal{V}'$. The total number of categories encoded is $\sum_{\mathcal{V}' \in \mathcal{V}} |\mathcal{V}'|$. (We could define a more usual design matrix by sequentially appending a column for each category of each predictor variable—that is what's implemented numerically. But the idea of $v$ within $\mathcal{V}'$ within $\mathcal{V}$ is needed when we come to the priors for the regression coefficients in equation 15.)
- $\beta^\theta_{\mathcal{V}'v}$ is the regression coefficient for the normal mixture model parameter $\theta$ (one of $p^{\text{pos}}, \mu^{\text{pos}}, \mu^{\text{neg}}, \sigma^{\text{pos}}, \sigma^{\text{neg}}$) associated with categorical variable $\mathcal{V}'$ taking the value $v$.
- $X^{\text{binary}}_{i,b} = \begin{cases} 1, & \text{if } i\text{'s value for binary predictor variable } b \text{ is TRUE}, \\ 0, & \text{otherwise} \end{cases}$
  This is the design matrix for binary predictor variables, used to predict seroprevalence only (not the mean or standard deviation of antibody level distributions). We treat such variables separately from the other predictor variables, because using random effects to estimate a distribution of values for different categories suffers from overparameterisation when there are only two categories. For these predictor variables we use fixed effects instead.
- $\beta^{\text{binary}}_b$ is the regression coefficient (the increase in seroprevalence on a logit scale) associated with binary predictor variable $b$ taking the value TRUE compared to when it is FALSE.

### 1.3 Specifying the model

Everywhere that we state something about the studied system (rather than statements of logic) as being so, it should really say that we *model* it as being so. We suppress this to avoid much repetition.

Assuming no difference in chemical reactions due to the different pipetting times, the different replicates of the same sample or calibrator have the same $x$, therefore there is no need for an $r$ index for $x$. Assuming no difference in chemical reactions and no mislabelling of samples, the same sample on different plates has a single $x$ (and we use a different $c$ index to describe the same dilution calibrator on different plates), therefore there is no need for an $l$ index for $x$. Thus the $\log_e$ of the antibody level for sample $i$ is $x_i$, and for calibrator $c$ it's $x_c$, regardless of replicate or plate.

Different replicates of the same sample or calibrator are exchangeable, following the same normal distribution, with both mean and standard deviation depending on both $x$ and $l$. The mean has the same dependence on $x$ for all samples and calibrators on plate $l$, namely a 4PL function with parameters $\mathbf{f}_{l\cdot}$. The standard deviation has a different dependence on $x$ for samples and calibrators, also 4PL, sharing the steepness and inflection point parameters with the mean's dependence on x, but with different lower and upper asymptotes: $\sigma_{\text{min}}^{\text{sam}}$ and $\sigma_{\text{max}}^{\text{sam}}$ for samples, $\sigma_{\text{min}}^{\text{cal}}$ and $\sigma_{\text{max}}^{\text{cal}}$ for calibrators. Different observations are conditionally independent of each other given all model parameters. We can state all the points of this paragraph mathematically thus:

$$y_{ilr}^{\text{sam}} \sim N(\mu_l(x_i), \sigma_l^{\text{sam}}(x_i)^2) \tag{1}$$

$$y_{clr}^{\text{cal}} \sim N(\mu_l(x_c), \sigma_l^{\text{cal}}(x_c)^2) \tag{2}$$

$$\mu_l(x) := \text{4PL}(x \mid \mathbf{f}_{l\cdot}) \tag{3}$$

$$\sigma_l^{\text{sam}}(x_i) := \text{4PL}(x_i \mid (f_{l,1}, \sigma_{\text{min}}^{\text{sam}}, \sigma_{\text{max}}^{\text{sam}}, f_{l,4})) \tag{4}$$

$$\sigma_l^{\text{cal}}(x_c) := \text{4PL}(x_c \mid (f_{l,1}, \sigma_{\text{min}}^{\text{cal}}, \sigma_{\text{max}}^{\text{cal}}, f_{l,4})) \tag{5}$$

$x_i$ is normally distributed with one mean and variance if $i$ is seropositive, another mean and variance if $i$ is seronegative, with the means and variances depending on $i$:

$$P(x_i \mid s_i, \mu_i^{\text{pos}}, \sigma_i^{\text{pos}}, \mu_i^{\text{neg}}, \sigma_i^{\text{neg}}) = s_i N(x_i \mid \mu_i^{\text{pos}}, (\sigma_i^{\text{pos}})^2) + (1 - s_i) N(x_i \mid \mu_i^{\text{neg}}, (\sigma_i^{\text{neg}})^2) \tag{6}$$

$s_i$ is boolean so is Bernoulli distributed:

$$s_i \sim \text{Ber}(p_i^{\text{pos}}) \tag{7}$$

$s_i$ is $i$'s serostatus; $p_i^{\text{pos}}$ is the seroprevalence of $i$'s subpopulation, defined at the most granular level based on all of the categorical predictor variables in $\mathcal{V}^{p^{\text{pos}}}$.

We can use equation 7 to marginalise over $s_i$ in equation 6:

$$P(x_i \mid p_i^{\text{pos}}) = \sum_{s_i} P(x_i \mid s_i) P(s_i \mid p_i^{\text{pos}}) \tag{8}$$

$$= p_i^{\text{pos}} N(x_i \mid \mu_i^{\text{pos}}, (\sigma_i^{\text{pos}})^2) + (1 - p_i^{\text{pos}}) N(x_i \mid \mu_i^{\text{neg}}, (\sigma_i^{\text{neg}})^2) \tag{9}$$

The right-hand side of equation 9 is identical to that of equation 6, except with $p_i^{\text{pos}}$ replacing $s_i$. Recalling that $s_i$ is either 0 or 1 whereas $p_i^{\text{pos}}$ is continuous between 0 and 1, in equation 6 exactly one of the two normal distributions contributes to the right-hand side, depending whether $s_i$ is 0 or 1, whereas in equation 9 both normal distributions contribute: this is a two-component normal mixture model.

The two-component normal mixture model contains five parameters, all of which we specify regression models for, allowing them to vary by individual. Each of the categorical predictor variables used for a given parameter contributes to it additively, using an appropriate link function (to keep

$0 \le p_i^{\text{pos}} \le 1$ and $\sigma_i^{\text{pos}}, \sigma_i^{\text{neg}} \ge 0$):

$$p_i^{\text{pos}} = \text{logistic}\left(\text{logit}(p^{\text{pos}}) + \sum_{\mathcal{V}' \in \mathcal{V}^{p^{\text{pos}}}} \sum_{v \in \mathcal{V}'} X_{i\mathcal{V}'v} \beta_{\mathcal{V}'v}^{p^{\text{pos}}} + \sum_b X_{i,b}^{\text{binary}} \beta_b^{\text{binary}}\right) \tag{10}$$

$$\mu_i^{\text{pos}} = \mu^{\text{pos}} + \sum_{\mathcal{V}' \in \mathcal{V}^{\mu^{\text{pos}}}} \sum_{v \in \mathcal{V}'} X_{i\mathcal{V}'v} \beta_{\mathcal{V}'v}^{\mu^{\text{pos}}} \tag{11}$$

$$\mu_i^{\text{neg}} = \mu^{\text{neg}} + \sum_{\mathcal{V}' \in \mathcal{V}^{\mu^{\text{neg}}}} \sum_{v \in \mathcal{V}'} X_{i\mathcal{V}'v} \beta_{\mathcal{V}'v}^{\mu^{\text{neg}}} \tag{12}$$

$$\sigma_i^{\text{pos}} = \exp\left(\log(\sigma^{\text{pos}}) + \sum_{\mathcal{V}' \in \mathcal{V}^{\sigma^{\text{pos}}}} \sum_{v \in \mathcal{V}'} X_{i\mathcal{V}'v} \beta_{\mathcal{V}'v}^{\sigma^{\text{pos}}}\right) \tag{13}$$

$$\sigma_i^{\text{neg}} = \exp\left(\log(\sigma^{\text{neg}}) + \sum_{\mathcal{V}' \in \mathcal{V}^{\sigma^{\text{neg}}}} \sum_{v \in \mathcal{V}'} X_{i\mathcal{V}'v} \beta_{\mathcal{V}'v}^{\sigma^{\text{neg}}}\right) \tag{14}$$

The regression coefficients for different categories of the same predictor variable, i.e. the different elements of the vector $\boldsymbol{\beta}_{\mathcal{V}'\cdot}^{\theta}$, share a distribution to partially pool their estimates. For $p^{\text{pos}}, \sigma^{\text{pos}}, \sigma^{\text{neg}}$ this is a simple normal distribution with mean zero and standard deviation $\sigma_{\mathcal{V}'}^{\theta}$:

$$\beta_{\mathcal{V}'v}^{\theta} \sim N(0, (\sigma_{\mathcal{V}'}^{\theta})^2), \qquad \text{for } \theta \in \{p^{\text{pos}}, \sigma^{\text{pos}}, \sigma^{\text{neg}}\} \tag{15}$$

For $\mu^{\text{neg}}$ and $\mu^{\text{pos}}$, we use truncated normal distributions:

$$\beta_{\mathcal{V}'v}^{\mu^{\text{neg}}} \sim N(0, (\sigma_{\mathcal{V}'}^{\mu^{\text{neg}}})^2)[-\infty, \tfrac{1}{2}(\mu^{\text{pos}} - \mu^{\text{neg}})] \tag{16}$$

$$\beta_{\mathcal{V}'v}^{\mu^{\text{pos}}} \sim N(0, (\sigma_{\mathcal{V}'}^{\mu^{\text{pos}}})^2)[\tfrac{1}{2}(\mu^{\text{neg}} - \mu^{\text{pos}}), \infty] \tag{17}$$

This is to make it impossible for any individual $\beta_{\mathcal{V}'v}^{\mu^{\text{pos}}}$ coefficient to become so negative, or any individual $\beta_{\mathcal{V}'v}^{\mu^{\text{neg}}}$ coefficient to become so positive, that when added onto $\mu^{\text{pos}}$ and $\mu^{\text{neg}}$ respectively (as in equations 11 and 12) the latter is greater than the former. See section 1.4 for more explanation.

Summarising these regression models in words, we use a separate random effect term for each categorical predictor variable, partially pooling estimates of the regression coeffecients for different categories of the same categorical predictor variable. For example, having seen the scale of variability in seroprevalence between several countries, we assume that a new country will fall roughly within the same scale of variability and apply a normal penalty to the probability of being outside that scale. We do not pool estimates across different predictor variables: the different values of $\sigma_{\mathcal{V}'}^{\theta}$ for different $\mathcal{V}'$ are given separate top-level priors, not modelled as coming from a distribution with parameters to be estimated. For example, we assume that the scale of variability in seroprevalence between countries is wholly uninformative of the scale of variability between age groups. We do not pool estimates for the same predictor variable across different parameters $\theta$ of the normal mixture model. For example, we assume that the scale of variability in seroprevalence ($\theta = p^{\text{pos}}$) between countries is wholly uninformative of the scale of variability in the mean antibody level of seropositives ($\theta = \mu^{\text{pos}}$) between countries.

The four-vector $\mathbf{f}_{l\cdot}$ is normally distributed over $\mathcal{L}$, with mean $\hat{\mathbf{f}}$ and 4x4 covariance matrix $\boldsymbol{\Sigma}$:

$$\mathbf{f}_{l\cdot} \sim N(\hat{\mathbf{f}}, \boldsymbol{\Sigma}) \tag{18}$$

In addition to random plate-level variability in the four-vector $\mathbf{f}_{l\cdot}$, a regression model can be specified for it. (We anticipate that this will typically not be of interest: variability in $\mathbf{f}_{l\cdot}$ between plates is typically a nuisance that we merely want to correct for, to ensure correct estimation of $x$ from $y$ on each plate, rather than something we want to understand systematically.) The regression model for $\mathbf{f}_{l\cdot}$ is similar to those for the five parameters of the normal mixture model for $x$, equations 10–14: each of a number of categorical variables contributes additively, with partial pooling of regression coefficients for different categories of the same categorical variable. No link function is required for $\mathbf{f}_{l\cdot}$ because it is unconstrained (c.f. a logit link function for $p_i^{\text{pos}}$ and a log link function for $\sigma_i^{\text{pos}}, \sigma_i^{\text{neg}}$). Because $\mathbf{f}_{l\cdot}$ is a four-vector, instead of the scalar regression models in equations 10–14, the normal distribution for different categories of the same predictor variable becomes a four-dimensional normal (c.f. scalar equation 15). For this we introduce new parameters for the four variances for each predictor variable, but re-use the dimensionless correlation matrix used for plate-level variation in equation 18.

#### 1.3.1 Accidentally blank wells

We have a variant of the main model that allows for the possibility of, and estimates the frequency of, some wells in plates accidentally containing no sample at all (a failure to pipette anything into the well). If this occurs the effective $x$ for that well is zero. This occurs independently with probability $p^{\text{blank}}$ in each well. Marginalising over whether this occurs, the distribution for $y$—equation 1—becomes a two-component normal mixture model:

$$y_{ilr}^{\text{sam}} \sim (1 - p^{\text{blank}}) N(\mu_l(x_i), \sigma_l^{\text{sam}}(x_i)^2) + p^{\text{blank}} N(\mu_l(0), \sigma_l^{\text{sam}}(0)^2) \quad (19)$$

$$= (1 - p^{\text{blank}}) N(\mu_l(x_i), \sigma_l^{\text{sam}}(x_i)^2) + p^{\text{blank}} N(f_{l,2}, (\sigma_{\text{min}}^{\text{sam}})^2) \quad (20)$$

### 1.4 Top-level priors

In section 1.3 we specified the priors for all lower-level parameters (‘random effects’, those parameters with prior distributions that depend on other parameters that vary, i.e. parameters that form part of the parameter space we explore during inference). We now clarify the priors for top-level parameters (‘fixed-effects’, those with prior distributions whose parameters (‘hyperparameters’) are kept fixed during inference).

For all except two of the top-level parameters we use uniform priors, with the upper and lower bounds chosen by the user at time of analysis (not hardcoded in advance). We parameterise $\mathbf{\Sigma}$ through a decomposition into a diagonal matrix of the marginal variances (the square roots of which have uniform priors) and a dimensionless correlation matrix (with an LKJ distribution as the prior). For $\mu^{\text{neg}}$ and $\mu^{\text{pos}}$, the dvsb user specifies upper and lower bounds as for all the other top-level parameters (except the aforementioned dimensionless correlation matrix). If these two ranges do not overlap, they are used to specify two uniform distributions in the intuitive way: $\mu^{\text{neg}} \sim \text{Unif}(\mu_{\text{min}}^{\text{neg}}, \mu_{\text{max}}^{\text{neg}})$, $\mu^{\text{pos}} \sim \text{Unif}(\mu_{\text{min}}^{\text{pos}}, \mu_{\text{max}}^{\text{pos}})$. If these ranges do overlap, then we demote $\mu^{\text{pos}}$ from a top-level parameter to a lower-level parameter, with the prior $\mu^{\text{pos}} \mid \mu^{\text{neg}} \sim \text{Unif}(\max(\mu_{\text{min}}^{\text{pos}}, \mu^{\text{neg}}), \mu_{\text{max}}^{\text{pos}})$, to constrain $\mu^{\text{pos}} > \mu^{\text{neg}}$. Without this constraint, the likelihood has a symmetry under a seropositive↔seronegative relabelling operation: exchanging these two categories with each other, i.e. identifying seropositive with seronegative and vice versa, we get back to the same likelihood. This would lead to a multimodal posterior, with a second mode having $\mu^{\text{pos}} < \mu^{\text{neg}}$, which damages efficient exploration of the posterior geometry and which we want to exclude on biological grounds (the $x$ distribution should be shifted to higher values for seropositives that for seronegatives). Equations 16 and 17 ensure that no *single* pair of $\beta_{\mathcal{V}'v}^{\mu^{\text{pos}}}$ and $\beta_{\mathcal{V}'v}^{\mu^{\text{neg}}}$ coefficients can reverse the order of $\mu^{\text{neg}}$ and $\mu^{\text{pos}}$. The sum over $\mathcal{V}'$ in equations 11 and 12, i.e. summing over *multiple* categorical variables to define a subpopulation via multiple different population partitions (such as both age and location), does in principle allow for $\mu_i^{\text{pos}} < \mu_i^{\text{neg}}$. However, we expect this parameter regime would be strongly penalised by the priors: four of the regression coefficients must be in the tails of their distributions. Furthermore, at time of writing, we have hardcoded $\mu^{\text{neg}}$ and $\mu^{\text{pos}}$ to accept no more than one categorical variable each in their regression models. This is the only way we could figure out how to code in Stan a truncated normal distribution for their regression coefficients and use a non-centered parameterisation for this distribution (making more orthogonal the relative sizes of the coefficients and their overall scale, for more efficient sampling).

### 1.5 Implications of the model for classifying individual samples

Bayes' theorem gives us the probability of seropositivity given a known $x_i$ and $p_i^{\text{pos}}$:

$$P(s_i = 1 \mid x_i, p_i^{\text{pos}}) = \frac{P(x_i \mid s_i = 1)P(s_i = 1 \mid p_i^{\text{pos}})}{P(x_i \mid p_i^{\text{pos}})} \tag{21}$$

$$= \frac{p_i^{\text{pos}} N(x_i \mid \mu_i^{\text{pos}}, (\sigma_i^{\text{pos}})^2)}{p_i^{\text{pos}} N(x_i \mid \mu_i^{\text{pos}}, (\sigma_i^{\text{pos}})^2) + (1 - p_i^{\text{pos}}) N(x_i \mid \mu_i^{\text{neg}}, (\sigma_i^{\text{neg}})^2)} \tag{22}$$

If we knew all model parameters exactly, we could evaluate the above expression exactly to determine the probability of being seropositive as a function of $x_i$ (for a given combination of predictor categories such as country and age group; the function is different for each different combination).

Since we don't know all model parameters exactly, we have uncertainty in $P(s_i = 1 | x_i, p_i^{\text{pos}})$, quantified by its posterior distribution. Thus the question "what is *the* probability that $i$ is seropositive?", though it sounds natural, obscures the fact that there is uncertainty (captured by the posterior distribution) in this probability. The most helpful interpretation of $P(s_i = 1 \mid x_i, p_i^{\text{pos}})$ is: if we were to observe one exact value of $x_i$ many times, what fraction of those observations would be seropositive. This framing clarifies that we would not expect to know this fraction with infinite precision from finite data. If the posterior for $P(s_i = 1 \mid x_i, p_i^{\text{pos}})$ is concentrated at 0 or 1 for a particular $i$, then we can be confident $i$ is seronegative or seropositive respectively (conditional on model correctness as always). If the posterior is concentrated at 0.5 to a much greater extent than the prior is, then the model has been able to precisely estimate the $x$ distibutions for negatives and positives, but this particular point lies right where the distributions run into each other: there is irreducible uncertainty in its classification. If the posterior is similar to the prior (which may well have a mean or median at 0.5), the model has learned little. Note the final two scenarios are distinct even though they might be used similarly for binary classification of a given sample.

A limitation of our statistical model applies generally to the use of a two-component normal distribution to model two processes (here, generation of log antibody levels for seronegatives and for seropositives) where one process should dominate at all low values and the other at all high values. The normal distribution has a pathology for this purpose: unless the two normals are constrained to have identical variances (which is generally unjustified), whichever normal has the larger variance will dominate the mixture at both asymptotically large and asymptotically small values. Consider equation 22 as a function of $x$, for any or all individuals in the same subpopulation and thus sharing values for the mixture model parameters $p_i^{\text{pos}}, \mu_i^{\text{pos}}, \sigma_i^{\text{pos}}, \mu_i^{\text{neg}}, \sigma_i^{\text{neg}}$. If $\sigma_i^{\text{pos}} > \sigma_i^{\text{neg}}$, equation 22 tends to 0 both as $x \to \infty$ and as $x \to -\infty$; if $\sigma_i^{\text{pos}} > \sigma_i^{\text{neg}}$, equation 22 tends to 1 in both limits. The desired behaviour is that this probability be monotonic between limits of 0 as $x \to -\infty$ and of 1 as $x \to \infty$. Achieving this would require a different and probably richer model than a normal mixture, such as a skew-normal mixture with a prior constraining the allowable parameters to enforce monotonocity. Currently relying only on the normal mixture, our model facilitates evaluation of the issue by plotting the above function. If it is monotonic over the range of x spanned by the data, the asymptotic pathology is irrelevant; we found this to be the case in our motivating real dataset.

Furthermore, slight deviations from monotonicity at the edges of the empirical x range do not jeopardise the model's application when the aim is obtaining a reasonable estimate of subpopulation seroprevalence, for which optimal classification of every individual separately is not necessary.

#### 1.6 Posterior retrodictive checks

We first recap how posterior retrodictive checks work in general. We retrodict the observed data $y$ given our estimation of its generative process, i.e. we reconstruct the distribution from which we believe it was drawn, via

$$P(y' \mid y) = \int_\theta P(y' \mid \theta) P(\theta \mid y) d\theta \tag{23}$$

$P(y' \mid y)$ is the distribution of hypothetical new data $y'$ given the observed data $y$, generated by the same process but with fresh draws of stochasticity; if we judge that $y$ appears to have been drawn from this distribution, we judge that the model fits the data. $\theta$ is the set of model parameters, $P(y' \mid \theta)$ is the sampling distribution for new data $y'$, and $P(\theta \mid y)$ is the posterior. The distribution for the full dataset, $P(y' \mid y)$, is typically an unwieldy object, but we can use it to calculate the distribution for whatever summary statistics we like, to check how the model fits different aspects of the data.

We facilitate and recommend three posterior retrodictive checks in our method. The first is for the $y$ values for each calibrator across all plates given their $x$ values, to check how well we model the $x \leftrightarrow y$ relationship across all plates. (One of the reported model outputs is the probability, for each plate, of the plate-specific deviation in $\mathbf{f}$ parameters and of the calibrator $y$ values observed given $\mathbf{f}$; plates with the lowest values for this are most suspect for model fit in this regard, and should be prioritised for this check.) The second is for the set of $y$ values from each sample, to check for any inconsistencies between multiple measurements from the same sample, and for any samples with extreme $y$ values that are in tension with the assumed underlying distribution for $x$ given our estimated $x \leftrightarrow y$ relationship. (One of the reported model outputs is the observation probability of every $y$ value conditional upon all parameters; the samples with the lowest values are most suspect in this regard, and should be prioritised for this check.) The third is the subpopulation-level distribution of $y$ values for all distinct subpopulations that are modelled as having different $x$ distributions. Assuming the $x \leftrightarrow y$ relationship has been modelled well, this checks how well we model the distribution of $x$ and its variability between subpopulations. An important nuance in this check is to recognise that using the full posterior $P(\theta \mid y)$ in equation 23 would resample only from the observational stochasticity associated with re-measuring the same set of samples from the same individuals, because the full set of parameters $\theta$ includes individual-specific parameters. A more stringent check is to consider resampling new individuals. We resample the parameters for each individual's $x$ given the estimated $x$ distribution for that individual's subpopulation, by modifying equation 23 to become

$$P(y' \mid y) = \int_{\theta_{\text{pop}}, \theta_{\text{ind}}} P(y' \mid \theta_{\text{ind}}) P(\theta_{\text{ind}} \mid \theta_{\text{pop}}) P(\theta_{\text{pop}} \mid y) d\theta_{\text{pop}} \theta_{\text{ind}} \tag{24}$$

where $\theta_{\text{ind}}$ and $\theta_{\text{pop}}$ are the sets of individual- and (sub)population-level parameters respectively. Using $P(\theta_{\text{ind}} \mid \theta_{\text{pop}})$ rather than $P(\theta_{\text{ind}} \mid \theta_{\text{pop}}, y)$ means that the product $P(\theta_{\text{ind}} \mid \theta_{\text{pop}}) P(\theta_{\text{pop}} \mid y)$ is not the posterior for $\theta_{\text{ind}}$, but is rather the predictive distribution for the parameters of newly sampled individuals given both sampling stochasticity and our posterior uncertainty in $\theta_{\text{pop}}$.

### 2 Supplementary Results

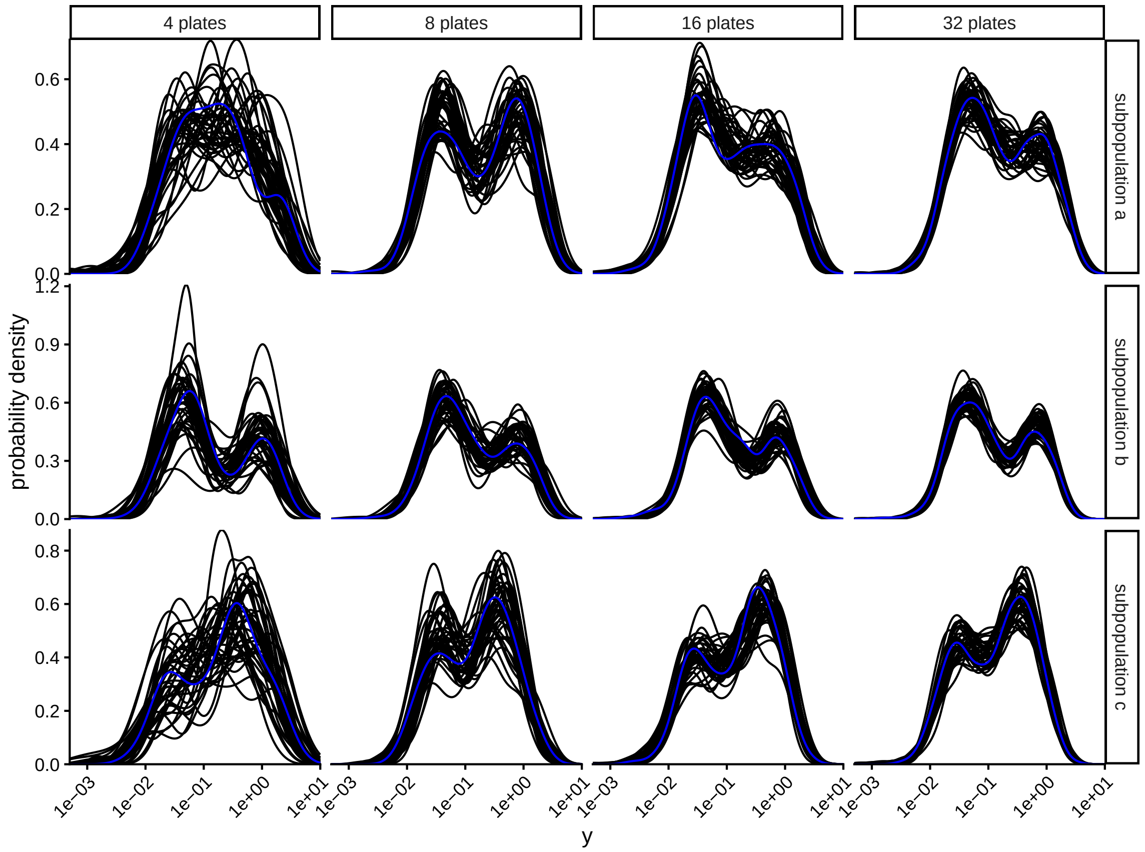


Supplementary Figure 1: A posterior retrodictive check for the results reported in main text regarding variable dataset size. Stratified by subpopulation, we show the distribution for $y$ (the serological assay proxy measurement for antibody level, such as ELISA OD value), as a kernel density estimate, with the empirical distribution in blue and the posterior fit to this distribution in black (one line per sample from the posterior, downsampled for clarity). We exclude the small fraction of negative y values to allow a logarithmic scale for clarity.

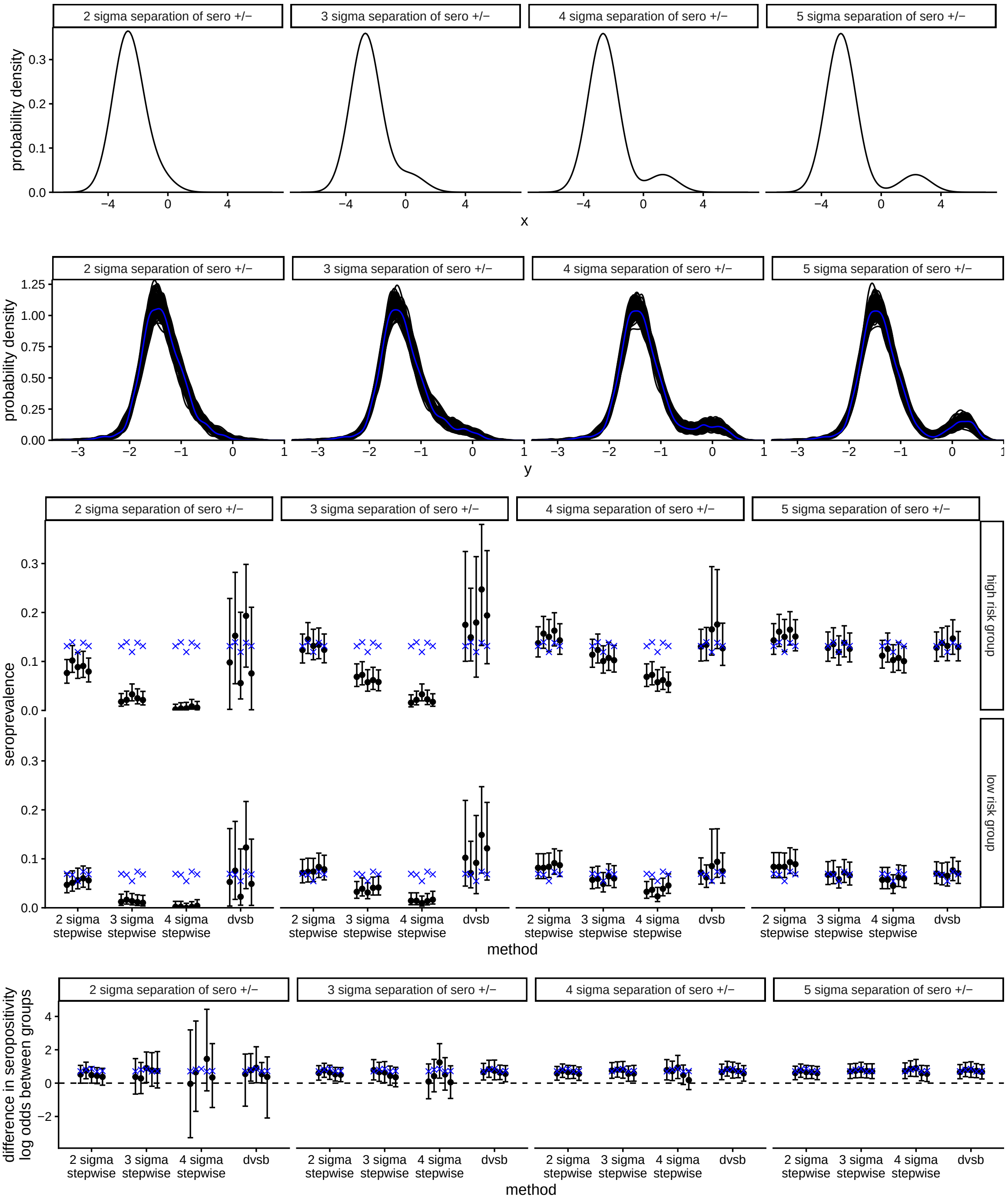


Supplementary Figure 2: As Figure 4 in main text, but for an overall seroprevalence of 10%.
